# Smartphone overuse and visual impairment in children and young adults: a systematic review and meta-analysis

**DOI:** 10.1101/2020.09.11.20192476

**Authors:** Jian Wang, Mei Li, Daqiao Zhu, Yang Cao

**Affiliations:** School of Nursing, Shanghai Jiao Tong University, 200025 Shanghai, China; School of Medical Sciences, Örebro University, 70182 Örebro, Sweden; Clinical Epidemiology and Biostatistics, School of Medical Sciences, Örebro University, 70182 Örebro, Sweden

**Keywords:** visual impairment, smartphone, mobile phone, overuse, child, young adult, systematic review, meta-analysis

## Abstract

**Background:** Smartphone overuse has been cited as a potentially modifiable risk factor that can result in visual impairment. However, associations between smartphone overuse and visual impairment have not been consistently reported.

**Objective:** The aim of this systematic review is to determine the association between smartphone overuse and visual impairment, including myopia, blurred vision, and poor vision, in children and young adults.

**Methods:** We conducted a systematic search in the Cochrane Library, Pubmed, EMBASE, Web of Science Core Collection, and ScienceDirect since the beginning of the databases to June 2020. Fourteen eligible studies (ten cross-sectional studies and four controlled trials) were identified, which included a total of 27110 subjects with mean ages ranging from 9.5 to 26.0 years. We used a random-effects model in the ten cross-sectional studies (*n* = 26962) and a fixed-effects model in the four controlled trials (*n* = 148) to combine odds ratios (OR) and effect sizes (ES). The *I*^2^ statistics was used to assess heterogeneity.

**Results:** A pooled OR of 1.05 (95% CI: 0.98, 1.13; *p* = 0.159) from cross-sectional studies suggests that smartphone overuse is not statistically significantly associated with myopia, poor vision, or blurred vision, however these visual impairments together are more apparent in children (OR = 1.06, 95% CI: 0.99, 1.14; p = 0.087) than in young adults (OR = 0.91, 95% CI: 0.57, 1.46; p = 0.707). In all the controlled trials, the smartphone overuse groups showed worse visual function scores compared with the less-use groups. The pooled ES is 0.76 (95% CI: 0.53, 0.99) and statistically significant (*p* < 0.001).

**Conclusions:** Our results indicate that longer smartphone use may increase the likelihood of ocular symptoms including myopia, asthenopia, and/or ocular surface disease, especially in children. Thus, regulating use time and restricting the prolonged use of smartphones may prevent ocular and visual symptoms. Further research on the patterns of use, with longer follow-up on the longitudinal associations will help inform detailed guidelines and recommendations for smartphone use in children and young adults.

## Introduction

The use of smartphones has been increasing rapidly since their introduction in the late 2000s [1]. In 2019, the global smartphone penetration had reached to about 41.5% of the global population [2]. Notably, the number of smartphone users in China was around 700 million in 2018, accounting for half of the Chinese population [3]. Data also showed that more than 80% of people in the United Kingdom owned or had ready access to a smartphone in 2019, with a significant increase from 50% in 2012 [4]. Furthermore, more than 90% of young people between 16 and 34 years old in the United Kingdom owned a smartphone [4].

With the continuous rise in youth’s digital media consumption, ocular problems have also seen a dramatic increase. A large population is suffering from visual impairment, especially in Asian countries, due to its rapidly increasing prevalence and younger age of onset [5–8]. It has been estimated that 49.8% (4.8 billion) and 9.8% (0.9 billion) of the global population will have myopia or high myopia by the year 2050 [9]. A recent study indicated that, about sixty years ago, only 10 – 20% of the Chinese population was shortsighted, but the percentage has been up to 90% of teenagers and young adults in 2015 [10]. Consistently, a school-based, retrospective, longitudinal cohort study (*n*=37424) found that the prevalence of myopia significantly increased from 56% in 2005 to 65% in 2015 [8].

Therefore, smartphone overuse among children and young adults has raised crucial concern in the society [11–13]. Several studies found an increased use of digital devices in children aged 2 – 11 years old [14, 15]. For example, a study of children aged 9 – 11 years old from 12 countries showed that 54.2% of the children exceeded proposed screen time guidelines (≤ 2 hours per day) [15]. Compared with older people, greater risks of the undesirable consequences in children and young adults have been reported because they have less self-control in smartphone use [11]. A cross-sectional study (*n* = 2639) indicated that 22.8% of teenagers were smartphone addiction users, which was related to hypertension [16]. Another research showed that the users of mobile devices spent > 20 hours weekly using email, text messages, and social networking services, indicating the heavy reliance on smartphones in their communication with other people [17]. Overall, smartphone overuse may result in significant harmful physical, psychological, and social consequences [18, 19].

Some experimental studies indicate that long time use of smartphone plays a key role in visual impairment, increasing the likelihood of poor vision [20–22]. For instance, a prospective clinical study (*n* = 50) showed that smartphone use for 4 hours resulted in a higher ocular surface disease index than those at baseline [20]. Kim et al found that the increase of ocular symptoms extended to the general population, especially in adolescents, after the expansion of smartphone use [23]. However, there are also studies that reported the lack of evidence for such association [24]. For example, a cross-sectional study (*n* = 1153) using stratified random cluster samples did not find a statistically significant association between smartphone use time and myopia [25]. Similarly, a research in Ireland (*n* = 418) indicated that smartphone use time was not a risk factor for myopia [26]. Also, Toh et al found that smartphone use time was associated with increased risk of visual symptoms (i.e. blurring of vision, dry eye), but decreased odds of myopia [27].

Although there has been increased concern about impaired vision by smartphone overuse, existing quantitative evidence about the relationships between excessive smartphone use and visual impairment is equivocal. It is essential to confirm and quantify whether excessive smartphone use may result in visual impairment, especially in children and young adults.

The aim of this study is to conduct a systematic review to summarize the existing evidence on the associations between smartphone overuse and visual impairment in children and young adults, which may further guide potential interventions to reduce the harmful impact of smartphone overuse on vision in the susceptible subpopulation group.

## Materials and Methods

### Data sources and search strategy

To carry out the systematic review and meta-analysis, we used a protocol that was constructed in line with the standard criteria, the Preferred Reporting Items for Systematic Reviews and Meta-Analysis (PRISMA) [28] and the Meta-analysis of Observational Studies in Epidemiology (MOOSE) [29].

A systematic search was carried out in Pubmed (the United States National Library of Medicine), Embase (Wolters Kluwer Ovid), Web of Science Core Collection (Clarivate Analytics), ScienceDirect (Elsevier), and Cochrane library (John Wiley & Sons, Ltd.) for observational and experimental studies that investigated smartphone overuse or addiction in children (age < 18 years) or young people (age < 40 years), and its associations with impaired visual function, such as myopia, poor vision, or blurred vision. To minimize publication bias, we also searched for additional studies in grey literature sources including Virtual Health Library (http://bvsalud.org/en/), NARCIS (https://www.narcis.nl/), Grey literature report (http://greylit.org/),and Open grey EU (http://opengrey.eu/). The search was limited in the publications published in English.

Free text and the Medical Subject Headings (MeSH) terms were used for the search, including:phone, smartphone, mobile/cell/cellular phone, electronic device, use, use time, screen time, overuse, addiction, eye, visual acuity, vision, vision screening, eyesight, myopia, myopic refraction, shortsighted/nearsighted/short sight, near sight, refraction errors, /ocular/health effect, optic, blind, ophthalmology, optometry, retina, ametropia/amblyopia symptom, visual assessment, and visual problem etc. (Supplemental material 1). We included all the observational studies and controlled trials (randomized or non-randomized), addressing smartphone use and visual impairment in human since the beginning of the databases to June 2020. Furthermore, manual retrieval was performed afterwards to ensure the inclusion of the latest literature.

### Inclusion and exclusion criteria

All observational and experimental studies were included if they have fulfilled the following criteria:

1. All original studies have examined the use of a smartphone (and/or mobile phone) and eyesight, including population-based longitudinal studies, cohort studies, case-control studies, cross-sectional studies, and controlled clinical trials;
2. The participants are children age ≤18 years or young people with age ≤ 40 years; the young adulthood in the study was defined as the developmental stage of those who are between 18 years old and 40 years old [30, 31];
3. The frequency or time of smartphone use (in minutes or hours, or per day or per week, et al.) have been reported;
4. The endpoint of interest is the incidence of visual impairment or decline, including myopia, poor vision, blurred vision, various visual function scores indicating impaired vision, and/or other unspecific visual impairments;
5. Vision measurements of the groups that may be used to calculate effect size (ES) of visual impairment, or odds ratio (OR) for the risk of visual impairment have been reported, as well as the associated 95% confidence interval (CI) or other data to estimate the variance or accuracy (such as standard error) were reported

Studies were excluded if they:

1. Are narrative reviews, editorial papers, commentaries, letters, or methodological papers;
2. Have evaluated visual function, or no reliable/relevant estimates for smartphone use;
3. Have no reference or control group included in the analysis;
4. Are animal studies.

### Data extraction

After the systematic search of the relevant articles in the databases, two investigators (J.W. and M.L.) embarked on screening and identification of potentially relevant abstracts independently. For any disagreements that occurred between the two investigators regarding the eligibility of a study, there was a thorough discussion or advice from an academic expert (Y.C.). Later, articles for selected abstracts were downloaded and data extracted by J.W. and Y.C. independently by use of a standardized form in Microsoft excel. Data extracted were compared and summarized to have one final document on which analysis was conducted. The information extracted included: name of the first author, year of publication, study design, duration of study, country that the study was conducted in, eyesight measurement, smartphone use time, smartphone use frequency, sample size, incidence cases with impaired vision, outcome ascertainment method, OR or ES and the associated 95% CI, and statistical analysis method used.

### Study quality assessment

The Joanna Briggs Institute (JBI) Critical Appraisal Checklist for Analytical Cross Sectional Studies, JBI Appraisal Checklist for Quasi-Experimental Studies, and JBI Critical Appraisal Checklist for Randomized Controlled Trials were used to assess the quality of the studies included in the meta-analysis [32]. J.W. and Y.C. assessed the quality of the articles independently then the final assessment was achieved upon discussion (Supplemental material 2, Tables S1 – S3).

### Statistical analysis

For studies that did not reported OR, OR was calculated using the numbers of cases with and without visual impairment of the reference/control group and overuse group. For studies that measured visual impairment using continuous variables, ES was calculated as the difference between the means divided by the pooled standard deviation (SD) [33]:

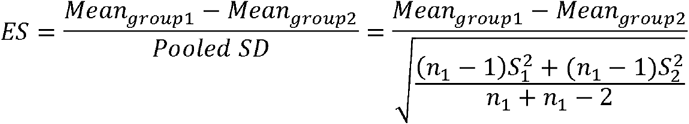

where *n*_1_, *S*_1_ and *n*_2_, *S*_2_ are sample size and standard deviation for group 1 and group 2, respectively.

A positive ES indicates a worse visual function. Heterogeneity of the included studies was investigated using *I*^2^ statistics[34]. *I*^2^ > 30% was considered moderate heterogeneity while *I*^2^ > 50% was considered substantial heterogeneity [35]. A p-value < 0.05 from the non-central chi-squared test for heterogeneity was considered statistically significant[36]. Contribution of each study to the heterogeneity and their influence on the pooled OR or ES was presented using Baujat plot [37]. The pooled OR with corresponding 95% CI were calculated using random effect models because heterogeneity presented among the studies, and presented using forest plot [38]. The possibility of publication bias was assessed by the combination of Egger’s test and visual inspection of funnel plot [39].

Subgroup analysis was performed for cross-sectional studies by outcome of visual impairment (myopia, poor vision, or blurred vision) and mean age of the subjects (children, mean age ≤ 18 years, or young people, mean age > 18 and ≤ 40 years). Leave-one-out (LOO) analysis to investigate the influence of a single study on the pooled effect was also performed as sensitivity analysis [40].

A two-sided p-value < 0.05 of the pooled estimates was considered statistically significant unless otherwise specified. All the analysis was performed in R 4.0.0 (the R Foundation for Statistical Computing, Vienna, Austria) using packages *meta* 4.12-0 [41] and *dmetar* 0.0.9000 [42].

## Results

### Characteristics of the studies

In total, 1961 articles were obtained from all the databases. After removing the duplicates, 1796 articles remained, and 121 of them were considered relevant for the meta-analysis after screening of the titles and abstracts. After screening full-text of the 121 articles downloaded, 14 articles met our inclusion criteria. These included ten cross-sectional studies and four controlled trials, which consisted of 27110 participants with mean ages ranging from 9.5 to 26.0 years. The flowchart of article searching and screening is shown in Figure 1. The ten cross-sectional studies addressed incidents of myopia [24–27, 43], blurred vision [44–46], poor vision and other unspecified visual impairments [23, 27, 44, 47]. In our analysis, those unspecified visual impairments were treated as poor vision. There are two studies [27, 44] that addressed two visual impairment outcomes, and each outcome was treated as a single study in the meta-analysis. The four studies that used the controlled trial design addressed ocular surface disease index score [20], asthenopia score [21], oculomotor function [48], and viewing distance [22]. A more detailed summary of characteristics of the included studies is reported in Table 1.

**Figure 1.**
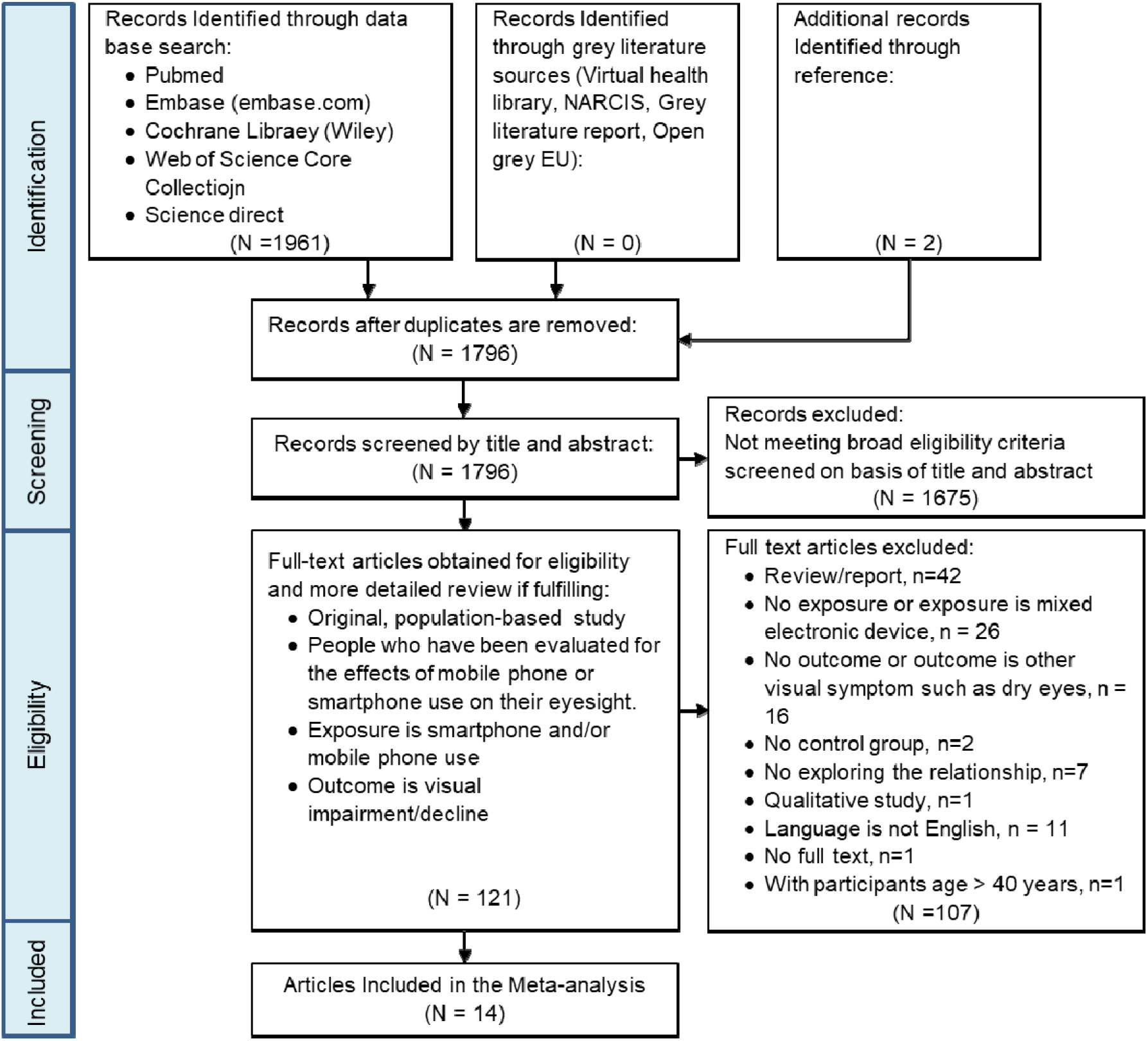
PRISMA flow diagram for screening and selection of articles on smartphone overuse and visual impairment in children and young adults

**Table 1.**
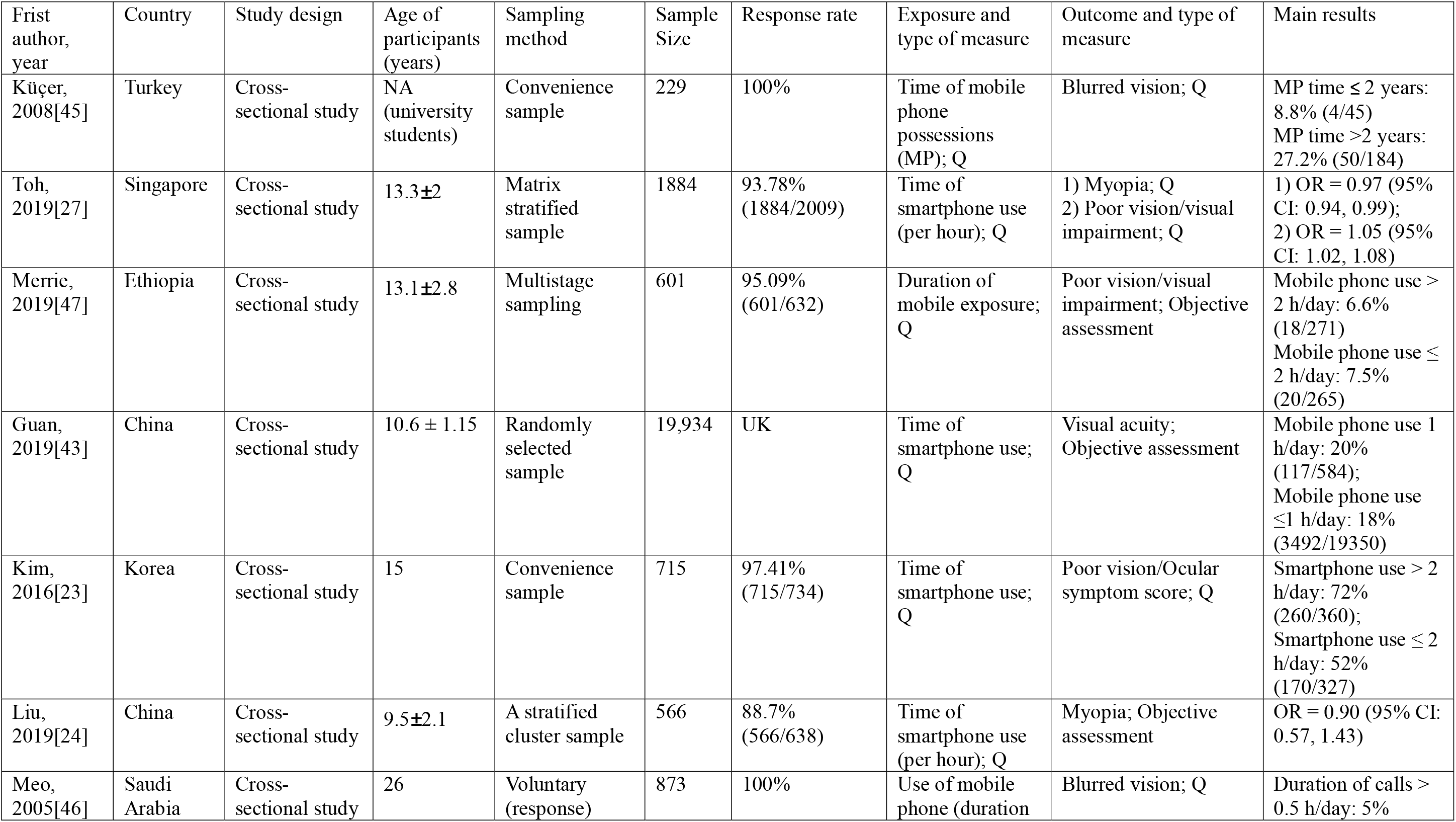
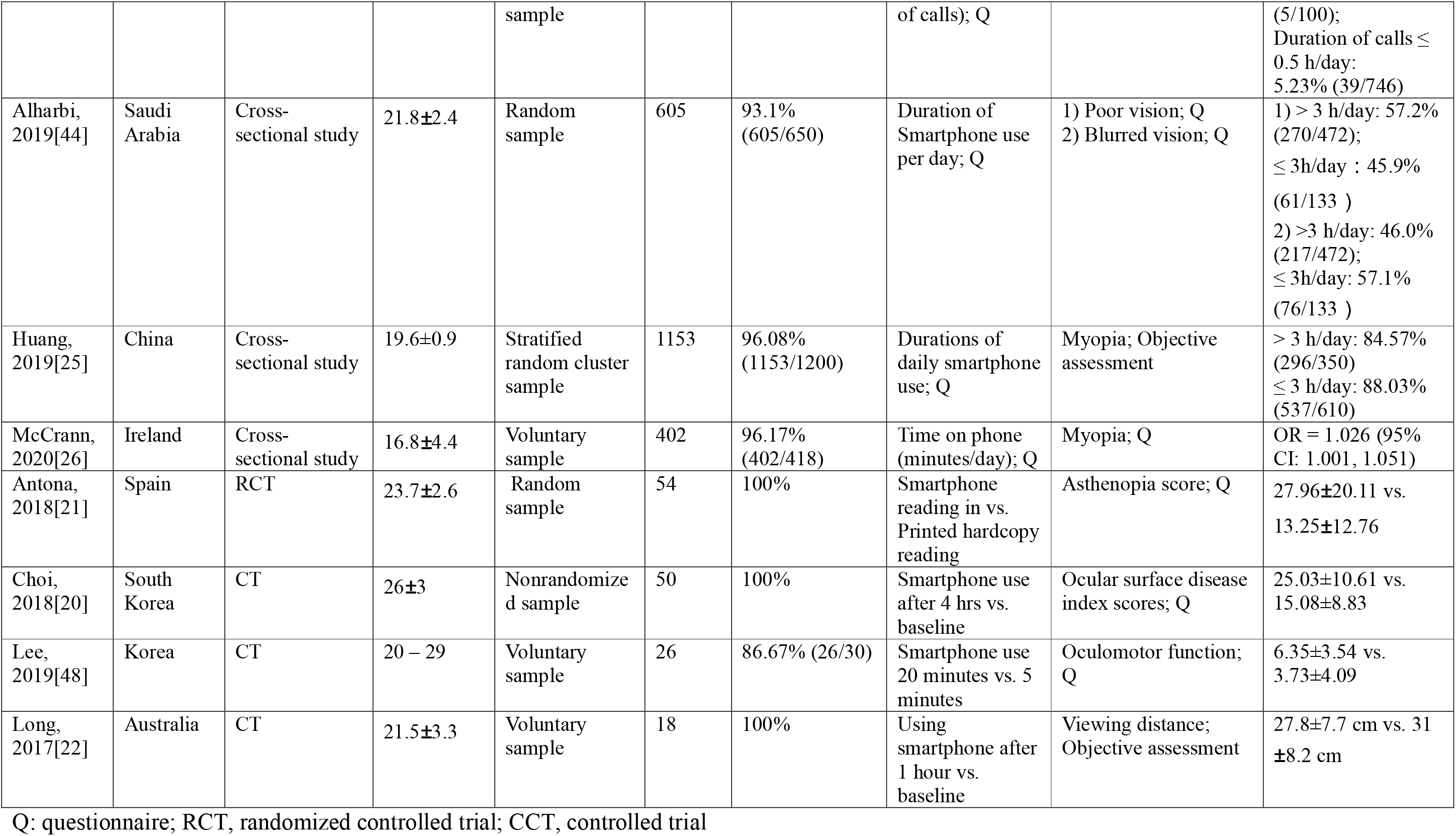
Characteristics of the included studies

### Association between smartphone overuse and incidence of visual impairment

The funnel plot of OR for the included cross-sectional studies appears symmetric (Figure 2). Although OR from two studies (Kim and Küçer) show a bit bias with other studies, no statistically significant publication bias was found by Egger’s test (p = 0.434).

**Figure 2.**
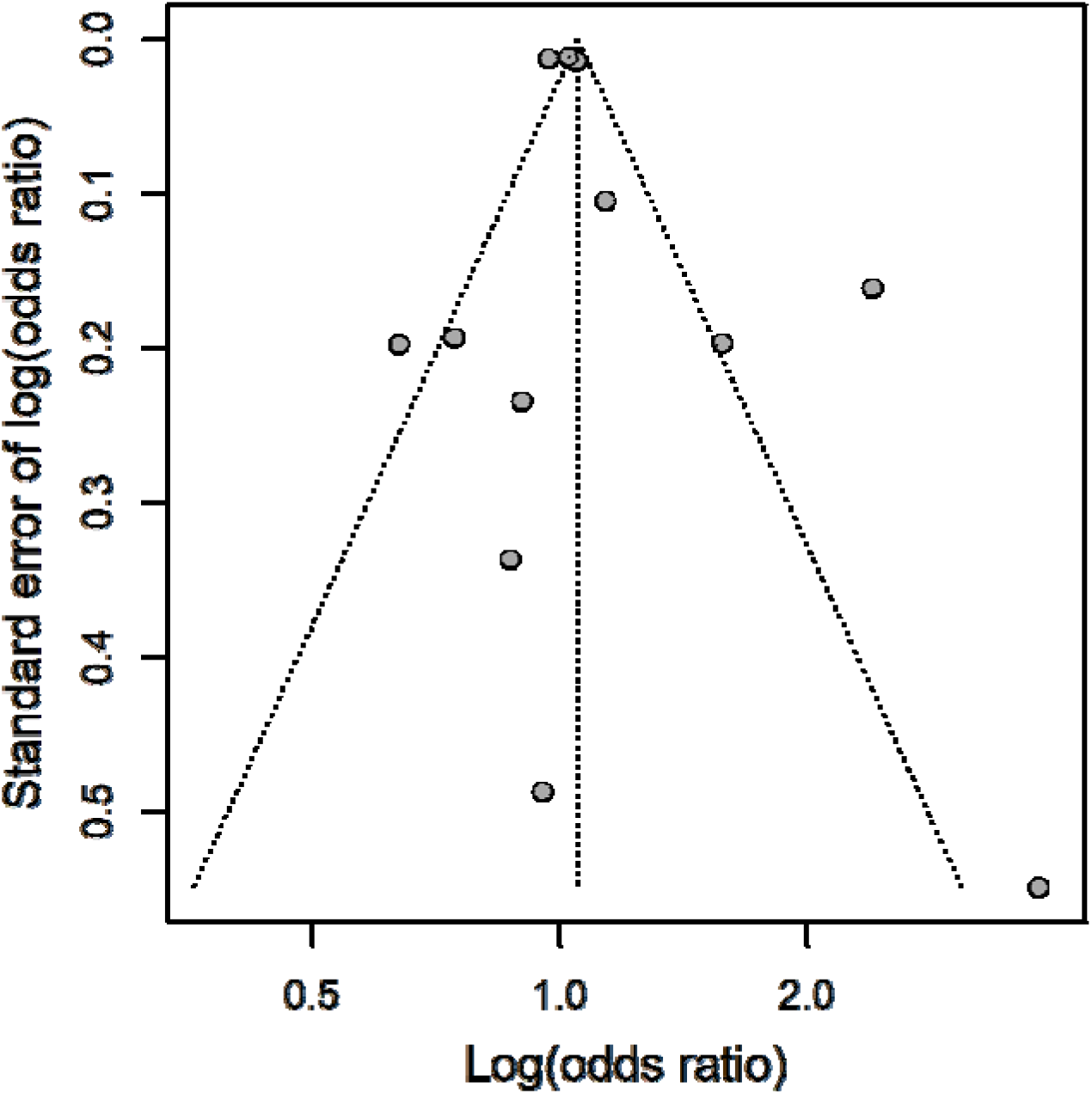
Funnel plot with pseudo 95% confidence limit for cross-sectional studies

Statistically significant heterogeneity was present among the ORs on visual impairment incidence (*I*^2^ = 84%, p< 0.001; Figure 3). Baujat plot indicates that Kim’s study contributed to the heterogeneity a lot while having few influences on the pooled OR (Figure 4). Overall, although the pooled OR shows that the odds of visual impairment is higher in the smartphone overuse group compared to the less-use group (OR = 1.05, 95% CI: 0.98, 1.13), the result is not statistically significant (p=0.159; Figure 3). None of the pooled ORs for specific visual impairment is statistically significant either in subgroup analysis. The pooled ORs for myopia, poor vision, and blurred vision are 1.00 (95% CI: 0.95, 1.05), 1.40 (95% CI: 0.87, 2.23), and 1.21 (0.44, 3.28), respectively (Figure 3). In neither age subgroups the pooled OR is statistically significant, which are 1.06 (95% CI: 0.99, 1.14; p = 0.087) for children and 0.91 (95% CI: 0.57, 1.46; p = 0.707) for young adults.

**Figure 3.**
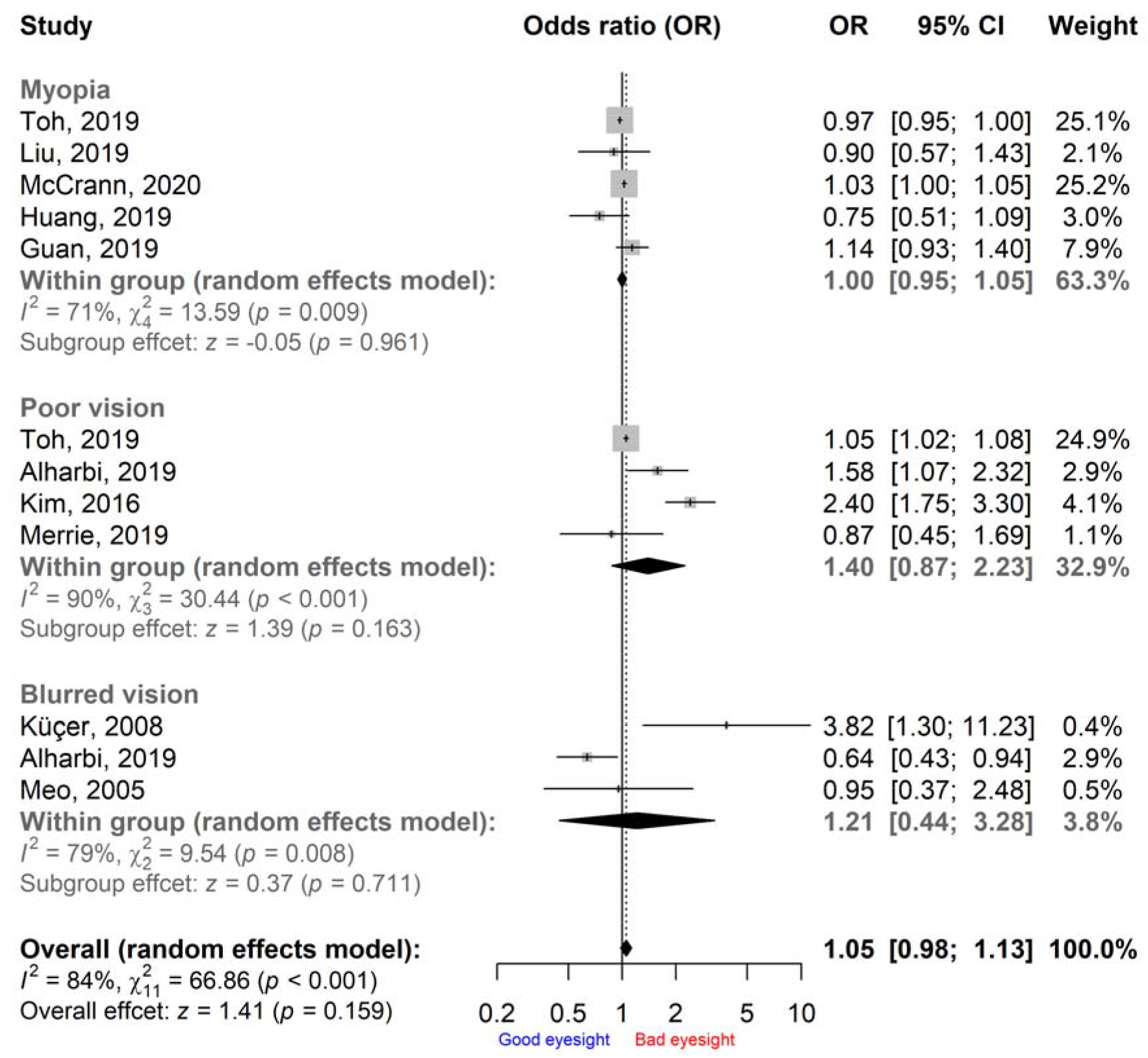
Pooled ORs of visual impairment in the smartphone overuse group compared to the less-use group

**Figure 4.**
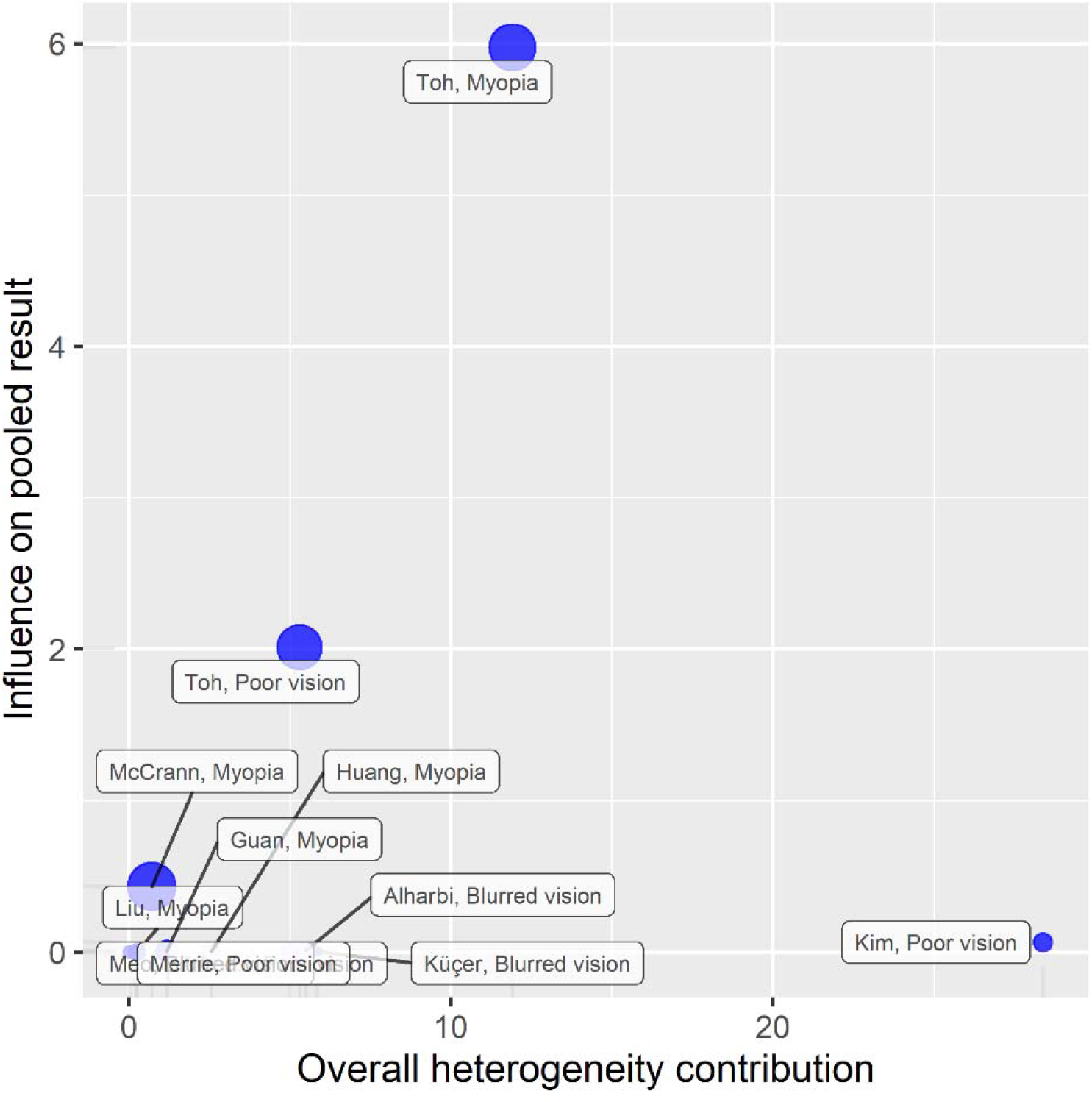
Baujat plot for cross-sectional studies

The LOO sensitivity indicates that ORs of visual impairment in the smartphone overuse group compared to the less-use group range from 1.02 to 1.09, however none of them is statistically significant (Figure 5).

**Figure 5.**
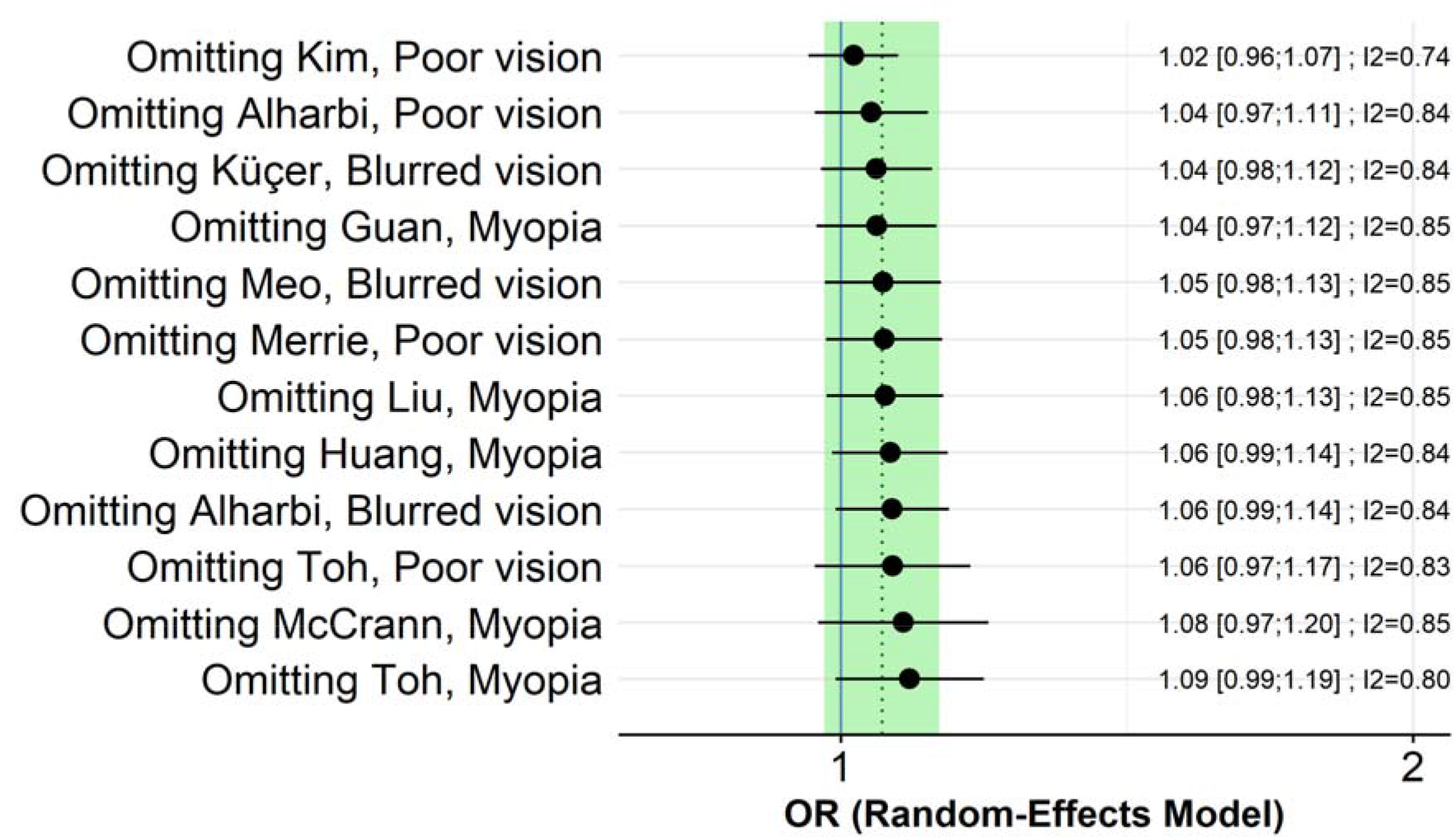
Pooled ORs of visual impairment in the smartphone overuse group compared to the less-use group from leave-one-out analysis

### Smartphone overuse associated worse visual function scores

The funnel plot of ES for the included controlled trial appears symmetric (Figure 6), and no statistically significant publication bias was found by Egger’s test (p = 0.066). No statistically significant heterogeneity was present among the ESs on visual impairment incidence (*I*^2^ = 0%, p=0.543; Figure 7).

**Figure 6.**
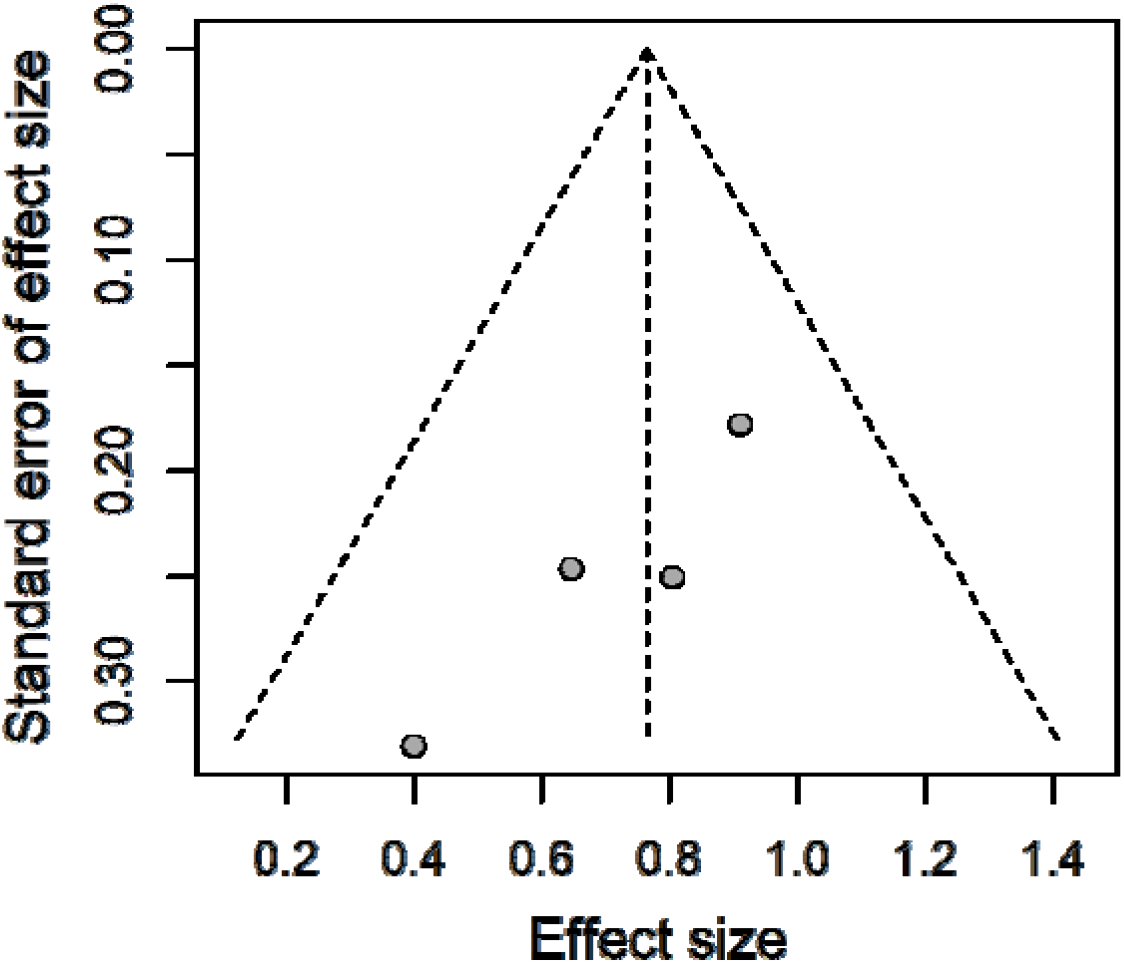
Funnel plot with pseudo 95% confidence limit for controlled trials

**Figure 7.**
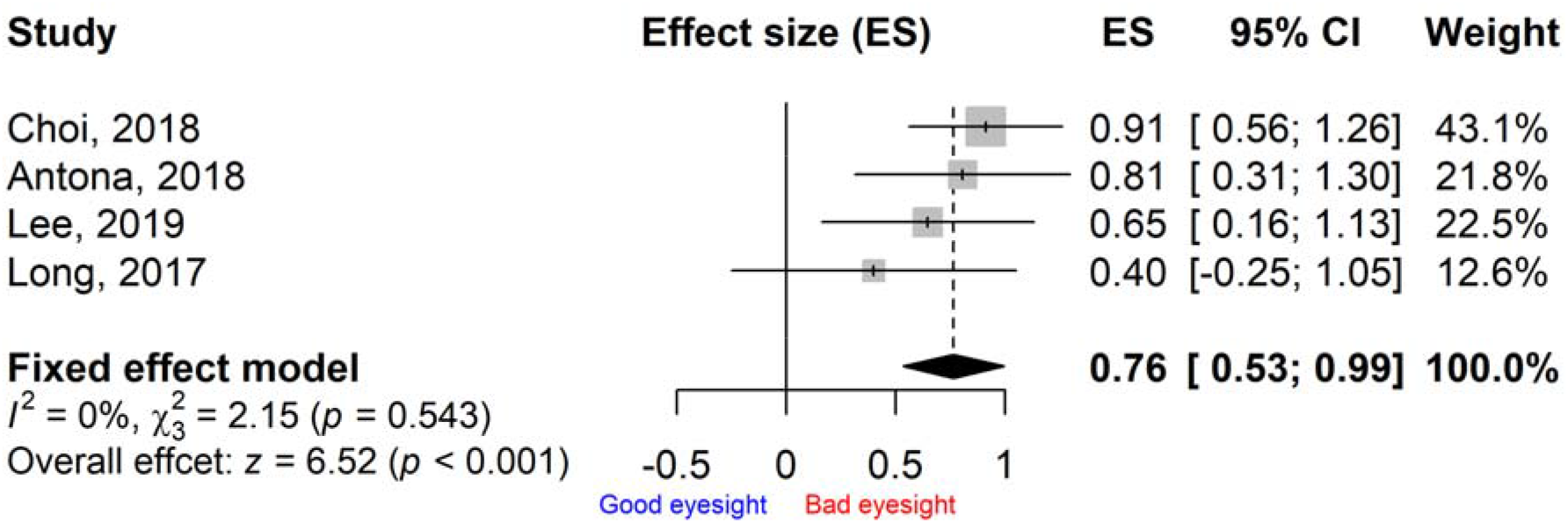
Pooled ES of visual function score in the smartphone overuse group compared to the less-use group

In all the controlled trials, the smartphone overuse group shows worse visual function scores than the less-use group, with ESs ranging from 0.40 to 0.91 (Figure 7). The pooled ES is 0.76 (95% CI: 0.53, 0.99) and statistically significant (p < 0.001), which means, compared to the less-use group, visual function score in the smartphone overuse group is 0.76 SD worse (Figure 7).

The LOO sensitivity indicates that the results are robust, with the ESs ranging from 0.65 to 0.82, and all the ESs are statistically significant (Figure 8).

**Figure 8.**
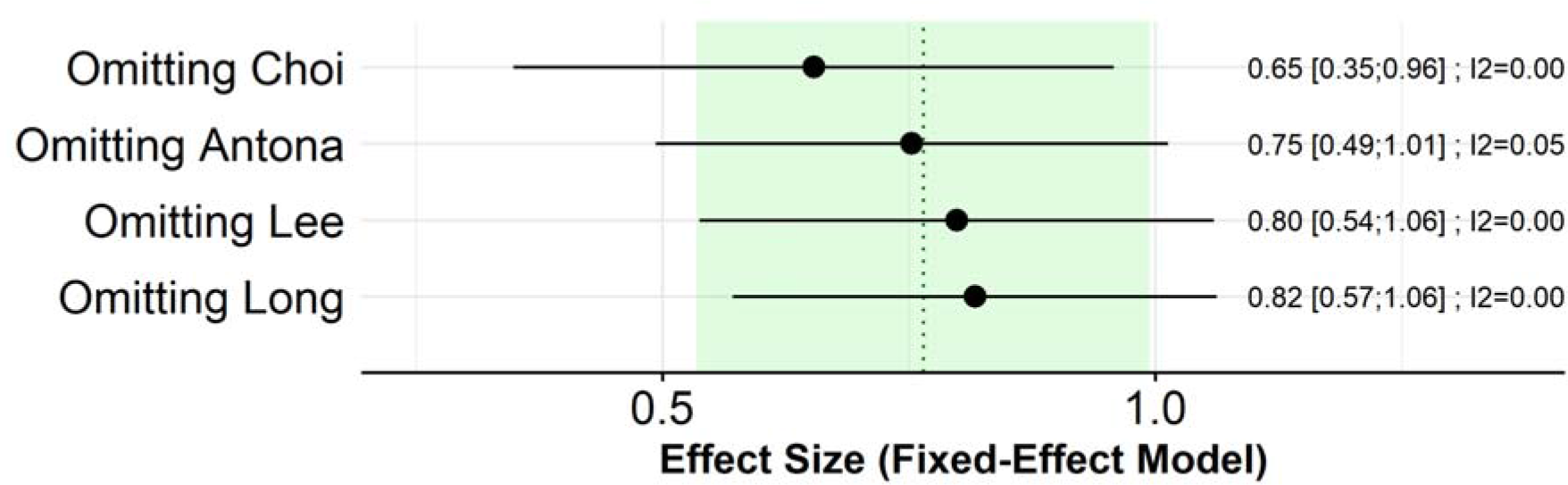
Pooled ESs of visual function score in the smartphone overuse group compared to the less-use group from leave-one-out analysis

## Discussion

The purpose of this systematic review and meta-analysis was to summarize currently available evidence with reference to the relationship between smartphone overuse and visual impairment in children and young adults. Nine out of fourteen studies found a significant association between smartphone overuse and visual impairment. Our pooled results show negative but not statistically significant associations (OR = 1.05, 95% CI: 0.98, 1.13) between smartphone overuse and myopia, blurred vision, or poor vision in the included cross-sectional studies. However, the adverse effect is more apparent in children (OR = 1.06, 95% CI: 0.99, 1.14) than in young adults (OR = 0.91, 95% CI: 0.57, 1.46). We also found that smartphone overuse may cause worse visual function than less use of smartphone in the included controlled trials (ES = 0.76, 95% CI: 0.53, 0.99). As the results are mixed, further studies are warranted. To our knowledge, this is the first systematic review that comprehensively summarized existing data on smartphone overuse and visual impairment in children and young adults.

No statistically significant association between smartphone overuse and visual impairment was found by pooling cross-sectional studies. The possible reasons might be as following.

Firstly, most of the existing studies in this systematic review were from Asia, representing the higher prevalence rates of visual impairment in these areas. However, evidence shows that the myopia prevalence in East Asia before the introduction of digital devices was already high [49]. Previous studies indicated that myopia prevalence increased more rapidly in people with more years of education and intensive schooling without particular exposure to screen devices [50–52]. For example, a study in Singapore found that myopia prevalence increased more rapidly in individuals that started elementary school after the 1980s [53]. Consistently, a study in Israel found that teenage boys who attended schools had much higher rates of myopia than students who spent less time on their books in the 1990s [49]. Therefore, education and intensive schooling may have a larger contribution to the increase in myopia prevalence. Recent studies have also extensively described the relationship between education and visual impairment [54]. Furthermore, the high prevalence of myopia from Taiwan was present in cohorts with low exposure to digital devices [52]. Thus, it is still debatable whether smartphone overuse would lead to a higher risk of myopia or other visual problems.

Secondly, most studies divided smartphone overuse as use time over 2 or 3 hours per day in our meta-analysis. However, some evidence showed that the time people spent on the digital screen was far longer [55–57], suggesting that people may use other electronic device. Other digital device overuse might also play an important role in visual impairment. Some studies have explored the relationships between digital screen time (e.g. computer, tablet, smartphone, or other handheld electronic screens) and visual impairment [56, 58–62]. For instance, a birth cohort study (n=5074) showed that increased computer use was associated with children’s myopia development [61]. Yang et al found that screen exposure was significantly and positively associated with preschool myopia [59], which is consistent with a cohort study [62]. However, the results of impacts of screen time on visual impairment were mixed. A recent systematic review showed that screen time was not statistically significantly associated with prevalent and incident myopia [58], which may largely support our pooled result of cross-sectional studies. Thus, the relationship needs to be further validated. Moreover, given differences in various digital devices use, some studies have compared the impacts of smartphones use with other digital devices on visual impairment [20, 24, 25, 27]. The results are inconsistent. For instance, Guan et al (*n* = 19934) found that prolonged (>60 minutes/day) computer usage and smartphone usage were both significantly associated with greater refractive error [43]. Nevertheless, Liu et al [24] and Huang et al [25] found that myopia in children was not associated with time spent using various electronic devices including smartphone, tablet and computers. Differently, a study with a representative sample of 1884 adolescents showed that smartphone use time was associated with increased risk of visual systems, but no significant association was found for tablet use [27]. A controlled trial (*n* = 50) indicated that smartphone use group got higher fatigue, burning, and dryness scores than the computer use group [20]. Although the existing research supported that smartphone use might cause worse vision than other digital devices, the results need further convincing evidence to be examined due to the low number of studies.

Thirdly, several studies have shown that technology use or screen time alone are of minimal risk to visual impairment, whilst more time spent outdoors is related to reduced risk of myopia and myopic progression [25, 63, 64]. However, some researchers believe that the increased use of digital devices may lead to more near work and less time spent outdoors, resulting in a substitution effect [58, 65]. For example, Dirani et al reported that the lack of adequate outdoor activity might be related to the rise in digital screen time [65]. To be specific, recent educational screen time might be a replacement of reading or writing, in addition to recreational screen time (e.g. computer or video games)[65]. For instance, smartphones are used by children mainly for playing games (29%) and watching videos (20%), but also for learning (19%) [66]. Thus, digital screen time might not be a causal factor, but maybe a substitute for a different type of near work. There is also some evidence that children 9 – 11 years old who spent < 2 hours playing on a computer were 1.98 times more likely to spend > 1 hour outside than those reporting two or more hours of computer use [67]. Although these results might reflect a trade-off between outdoor time and digital screen time, with screen time being a proxy of indoor time, there is no evidence to confirm this substitution effect. Thus, further studies in this field are warranted.

Besides the findings in the cross-sectional studies, we also found that smartphone overuse group presented worse visual function scores than the less-use group in all the included controlled trials and the pooled result. Biologically, the effects of smartphones on ocular symptoms can be explained by two types of electromagnetic fields (EMFs); extremely low-frequency EMFs and radiofrequency (RF) electromagnetic radiation (EMR) [68, 69]. The intensity of radiation from mobile phones is relatively low with a specific absorption rate < 4 W/kg [68, 70]. However, it has been reported that adverse effects, such as DNA damage and thickening of the cornea, occurs even at a specific absorption rate lower than 4 W/kg [68, 71, 72]. The local specific absorption rate has been shown higher in tissues at a younger age, suggesting higher susceptibility of adolescents to smartphones [73]. The EMFs generated by smartphones may interact with the tissues of the eyes [69, 74], which may cause apoptosis, cataract formation, edema, endothelial cell loss, inflammatory responses, and neurological effects [68, 70, 75, 76]. RF EMR may affect the body thermally and non-thermally [77], which may result in oxidative stress in the cornea and the lens [70]. These effects by EMFs and RF EMR on the eyes, especially on the cornea and the lens, could suggest why ocular symptoms such as blurring, redness, visual disturbance, inflammation and lacrimation increased when exposed to smartphones. Although experimental studies may make causal inferences, our result needs to be further confirmed due to limited number of the existing studies.

Regarding the association between smartphone overuse and myopia examined in the cross-sectional studies, multiple ocular symptoms found in the experimental studies do not mean pathological changes in the eyes like myopia. Few longitudinal cohort studies have examined the impacts of screen exposure on myopia, but the results are inconsistent [62, 78]. To our knowledge, there are no experimental or longitudinal studies detecting the impacts of the smartphone overuse on myopia. Considering longitudinal cohort studies establishing the temporal sequence of prior exposure to environmental factors, further research may use this study design to examine whether smartphone overuse may increase the risk of developing myopia.

In addition, the heterogeneity is high in the meta-analysis of included cross-sectional studies. First of all, a large number of studies have proved potential risk factors that may result in visual impairment, which included both genetic and environmental ones [20, 26, 52, 58] (e.g. age [26], education and occupation [58], outdoor activity [20, 58] and parental myopia [20]). However, some studies did not include these variables in the multivariate analysis, which might result in inconsistent findings and might further affect the individual effect estimates and the pooled OR. Secondly, some studies only used univariate analysis to investigate the associations between smartphone use time and visual impairment [43, 65], which might hinder the exploration of their interrelationships. Thirdly, the assessment of the outcome was inconsistent. For example, some studies used a self-reported questionnaire to identify myopia [26, 27], while some other used objective assessment [24, 25]. Furthermore, the division of smartphone overuse was inconsistent, which may preclude us from determining their significant relationships. A guideline advised limiting recreational screen time to no more than 2 hours per day [79]. Further study needs to use a broadly recognized standard to define smartphone overuse.

There are also other limitations need to be addressed. All included studies used self-reported questionnaire to evaluate smartphone use time. Participants in the included experimental studies also mostly reported their visual function using questionnaires. The questionnaires may be a potential source of error due to inaccurate reporting or recall bias of participants. Further research should adopt objective instruments to measure smartphone use time, and visual acuity (VA) screening to examine visual function. Furthermore, generalization of the results should be done with caution due to low number of studies included in each meta-analysis. Limiting the review to studies reported in English may also result in non-reporting of studies published in other languages. Nevertheless, our review has included rigorous methodological procedures to obtain and pool data from 27110 children and young adults. We also adopted a wide range of search terms to retrieve all potential articles published in English, including grey literature, which might help to reduce the publication bias in the combination.

## Conclusions

Overall, current evidence suggests that the result of smartphone overuse and visual impairment in children and young adults are mixed. Although the statistically significantly negative association between smartphone overuse and visual impairment in the meta-analysis was only confirmed in controlled trials but not in cross-sectional studies, the adverse effect of smart overuse on visual functions is apparent in children. However, the relationships need to be further verified. Further research on the patterns of use, with longer follow-up on the longitudinal associations, and the exact mechanisms behind these associations will help inform detailed guidelines for smartphone use in children and young adults. In addition, understanding the factors of smartphone overuse that account for the risk of ocular symptoms could help the growing population of smartphone users, especially children and young adults, to use smartphones in a healthier manner.

## Data Availability

The data used in the meta-analysis are available on request from the corresponding author.

## Author Contributions

Conceptualization, Daqiao Zhu, Mei Li and Yang Cao; Data curation, Jian Wang, Mei Li and Yang Cao; Formal analysis, Jian Wang and Yang Cao; Investigation, Jian Wang, Daqiao Zhu and Mei Li; Methodology, Daqiao Zhu and Yang Cao; Project administration, Yang Cao; Software, Yang Cao; Supervision, Daqiao Zhu and Yang Cao; Validation, Jian Wang and Yang Cao; Visualization, Yang Cao; Writing – original draft, Jian Wang and Yang Cao; Writing – review & editing, Jian Wang, Daqiao Zhu, Mei Li and Yang Cao.

## Data availability

The data used in the meta-analysis are available on request from the corresponding author.

## Abbreviations

CI: confidence interval
EMF: electromagnetic field
EMR: electromagnetic radiation
ES: effect size
JBI: Joanna Briggs Institute
LOO: leave-one-out
MeSH: Medical Subject Headings
MOOSE: Meta-analysis of Observational Studies in Epidemiology
OR: odds ratio
PRISMA: Preferred Reporting Items for Systematic Reviews and Meta-Analysis
Q: questionnaire
RF: radiofrequency
SD: standard deviation
VA: visual acuity

## References

1. Böhm S, editor An analysis on country-specific characteristics influencing smartphone usage. Proceedings of “The First International Conference on Multidisciplinary in Management”, Bangkok; 2015.

2. Ertemel AV, Ari E. A Marketing Approach to a Psychological Problem: Problematic Smartphone Use on Adolescents. Int J Env Res Pub He. 2020;17(7). doi: ARTN 2471 10.3390/ijerph17072471. PubMed PMID: WOS:000530763300303.

3. Zhai XY, Ye M, Wang C, Gu Q, Huang T, Wang K, et al. Associations among physical activity and smartphone use with perceived stress and sleep quality of Chinese college students. Ment Health Phys Act. 2020;18. doi: ARTN 100323 10.1016/j.mhpa.2020.100323. PubMed PMID: WOS:000533648100010.

4. Csibi S, Griffiths MD, Demetrovics Z, Szabo A. Analysis of Problematic Smartphone Use Across Different Age Groups within the ‘Components Model of Addiction’. International Journal of Mental Health and Addiction. 2019:1–16.

5. Li L, Zhong H, Li J, Li CR, Pan CW. Incidence of myopia and biometric characteristics of premyopic eyes among Chinese children and adolescents. Bmc Ophthalmol. 2018;18. doi: ARTN 178 10.1186/s12886-018-0836-9. PubMed PMID: WOS:000439440000002.

6. Wang SK, Guo Y, Liao C, Chen Y, Su G, Zhang G, et al. Incidence of and Factors Associated With Myopia and High Myopia in Chinese Children, Based on Refraction Without Cycloplegia. JAMA Ophthalmol. 2018;136(9):1017–24. Epub 2018/07/07. doi: 10.1001/jamaophthalmol.2018.2658. PubMed PMID: 29978185; PubMed Central PMCID: PMCPMC6142978.

7. Grzybowski A, Kanclerz P, Tsubota K, Lanca C, Saw SM. A review on the epidemiology of myopia in school children worldwide. Bmc Ophthalmol. 2020;20(1):27. Epub 2020/01/16. doi: 10.1186/s12886-019-1220-0. PubMed PMID: 31937276; PubMed Central PMCID: PMCPMC6961361.

8. Li Y, Liu J, Qi P. The increasing prevalence of myopia in junior high school students in the Haidian District of Beijing, he increasing prevalence of myopia in junior high school students in the Haidian District of Beijing China: a 10-year population-based survey. Bmc Ophthalmol. 2017;17(1):88. Epub 2017/06/14. doi: 10.1186/s12886-017-0483-6. PubMed PMID: 28606071; PubMed Central PMCID: PMCPMC5468969.

9. Holden BA, Fricke TR, Wilson DA, Jong M, Naidoo KS, Sankaridurg P, et al. Global Prevalence of Myopia and High Myopia and Temporal Trends from 2000 through 2050. Ophthalmology. 2016;123(5):1036–42. doi: 10.1016/j.ophtha.2016.01.006. PubMed PMID: WOS:000375942300025.

10. Dolgin E. The myopia boom. Nature. 2015;519(7543):276–8. Epub 2015/03/20. doi: 10.1038/519276a. PubMed PMID: 25788077.

11. Choi SW, Kim DJ, Choi JS, Ahn H, Choi EJ, Song WY, et al. Comparison of risk and protective factors associated with smartphone addiction and Internet addiction. J Behav Addict. 2015;4(4):308–14. Epub 2015/12/23. doi: 10.1556/2006.4.2015.043. PubMed PMID: 26690626; PubMed Central PMCID: PMCPMC4712765.

12. Liu CH, Lin SH, Pan YC, Lin YH. Smartphone gaming and frequent use pattern associated with smartphone addiction. Medicine. 2016;95(28). doi: ARTN e4068 10.1097/MD.0000000000004068. PubMed PMID: WOS:000380767200014.

13. Haug S, Castro RP, Kwon M, Filler A, Kowatsch T, Schaub MP. Smartphone use and smartphone addiction among young people in Switzerland. Journal of Behavioral Addictions. 2015;4(4):299–307. doi: 10.1556/2006.4.2015.037. PubMed PMID: WOS:000366946300011.

14. Bernard JY, Padmapriya N, Chen B, Cai S, Tan KH, Yap F, et al. Predictors of screen viewing time in young Singaporean children: the GUSTO cohort. Int J Behav Nutr Phys Act. 2017;14(1):112. Epub 2017/09/06. doi: 10.1186/s12966-017-0562-3. PubMed PMID: 28870219; PubMed Central PMCID: PMCPMC5584344.

15. LeBlanc AG, Katzmarzyk PT, Barreira TV, Broyles ST, Chaput JP, Church TS, et al. Correlates of Total Sedentary Time and Screen Time in 9–11 Year-Old Children around the World: The International Study of Childhood Obesity, Lifestyle and the Environment. PLoS One. 2015;10(6):e0129622. Epub 2015/06/13. doi: 10.1371/journal.pone.0129622. PubMed PMID: 26068231; PubMed Central PMCID: PMCPMC4465981.

16. Zou Y, Xia N, Zou Y, Chen Z, Wen Y. Smartphone addiction may be associated with adolescent hypertension: a cross-sectional study among junior school students in China. BMC Pediatr. 2019;19(1):310. Epub 2019/09/06. doi: 10.1186/s12887-019-1699-9. PubMed PMID: 31484568; PubMed Central PMCID: PMCPMC6724312.

17. Lee S, Kang H, Shin G. Head flexion angle while using a smartphone. Ergonomics. 2015;58(2):220–6. Epub 2014/10/18. doi: 10.1080/00140139.2014.967311. PubMed PMID: 25323467.

18. van Deursen AJAM, Bolle CL, Hegner SM, Kommers PAM. Modeling habitual and addictive smartphone behavior The role of smartphone usage types, emotional intelligence, social stress, self-regulation, age, and gender. Comput Hum Behav. 2015;45:411–20. doi: 10.1016/j.chb.2014.12.039. PubMed PMID: WOS:000350081500036.

19. Pan CW, Qiu QX, Qian DJ, Hu DN, Li J, Saw SM, et al. Iris colour in relation to myopia among Chinese school-aged children. Ophthal Physl Opt. 2018;38(1):48–55. doi: 10.1111/opo.12427. PubMed PMID: WOS:000418554600005.

20. Choi JH, Li Y, Kim SH, Jin R, Kim YH, Choi W, et al. The influences of smartphone use on the status of the tear film and ocular surface. Plos One. 2018;13(10). doi: ARTN e0206541 10.1371/journal.pone.0206541. PubMed PMID: WOS:000448823700128.

21. Antona B, Barrio AR, Gasco A, Pinar A, Gonzalez-Perez M, Puell MC. Symptoms associated with reading from a smartphone in conditions of light and dark. Appl Ergon. 2018;68:12–7. doi: 10.1016/j.apergo.2017.10.014. PubMed PMID: WOS:000426224900002.

22. Long J, Cheung R, Duong S, Paynter R, Asper L. Viewing distance and eyestrain symptoms with prolonged viewing of smartphones. Clin Exp Optom. 2017;100(2):133–7. doi:10.1111/cxo.12453. PubMed PMID: WOS:000395744100004.

23. Kim J, Hwang Y, Kang S, Kim M, Kim TS, Kim J, et al. Association between Exposure to Smartphones and Ocular Health in Adolescents. Ophthalmic Epidemiol. 2016;23(4):269–76. Epub 2016/06/03. doi: 10.3109/09286586.2015.1136652. PubMed PMID: 27254040.

24. Liu SX, Ye S, Xi W, Zhang X. Electronic devices and myopic refraction among children aged 6–14 years in urban areas of Tianjin, lectronic devices and myopic refraction among children aged 6–14 years in urban areas of Tianjin China. Ophthal Physl Opt. 2019;39(4):282–93. doi: 10.1111/opo.12620. PubMed PMID: WOS:000472766900007.

25. Huang LM, Kawasaki H, Liu YQ, Wang ZL. The prevalence of myopia and the factors associated with it among university students in Nanjing: A cross-sectional study. Medicine. 2019;98(10). doi: ARTN e14777 10.1097/MD.0000000000014777. PubMed PMID: WOS:000462570100055.

26. McCrann S, Loughman J, Butler JS, Paudel N, Flitcroft DI. Smartphone use as a possible risk factor for myopia. Clin Exp Optom. 2020. doi: 10.1111/cxo.13092. PubMed PMID: WOS:000535200000001.

27. Toh SH, Coenen P, Howie EK, Mukherjee S, Mackey DA, Straker LM. Mobile touch screen device use and associations with musculoskeletal symptoms and visual health in a nationally representative sample of Singaporean adolescents. Ergonomics. 2019;62(6):778–93. doi: 10.1080/00140139.2018.1562107. PubMed PMID: WOS:000470461000001.

28. Moher D, Liberati A, Tetzlaff J, Altman DG, Group P. Preferred reporting items for systematic reviews and meta-analyses: the PRISMA statement. Int J Surg. 2010;8(5):336–41. Epub 2010/02/23. doi: 10.1016/j.ijsu.2010.02.007. PubMed PMID: 20171303.

29. Stroup DF, Berlin JA, Morton SC, Olkin I, Williamson GD, Rennie D, et al. Meta-analysis of observational studies in epidemiology: a proposal for reporting. Meta-analysis Of Observational Studies in Epidemiology (MOOSE) group. JAMA. 2000;283(15):2008–12. Epub 2000/05/02. doi: 10.1001/jama.283.15.2008. PubMed PMID: 10789670.

30. Erikson EH, Erikson JM. The life cycle completed (extended version): WW Norton & Company; 1998.

31. Zastrow C, Kirst-Ashman K. Understanding human behavior and the social environment: Cengage Learning; 2006.

32. Joanna Briggs Institute. JBI Critical Appraisal Tools Adelaide, Australia: Joanna Briggs institute; 2017 [cited 2020 June 4]. Available from: https://joannabriggs.org/ebp/critical_appraisal_tools.

33. Rosnow RL, Rosenthal R. Computing contrasts, effect sizes, and counternulls on other people's published data: General procedures for research consumers. Psychol Methods.1996;1(4):331–40. doi: Doi 10.1037/1082-989x.1.4.331. PubMed PMID: WOS:A1996VW15400001.

34. Higgins JP, Thompson SG, Deeks JJ, Altman DG. Measuring inconsistency in meta-analyses. BMJ. 2003;327(7414):557–60. Epub 2003/09/06. doi: 10.1136/bmj.327.7414.557. PubMed PMID: 12958120; PubMed Central PMCID: PMCPMC192859.

35. Higgins J, Green S. Cochrane handbook for systematic reviews of interventions Version 5.1.0. London: The Cochrane Collaboration; 2018.

36. Higgins JP, Thompson SG. Quantifying heterogeneity in a meta-analysis. Stat Med. 2002;21(11):1539–58. Epub 2002/07/12. doi: 10.1002/sim.1186. PubMed PMID: 12111919.

37. Baujat B, Mahe C, Pignon JP, Hill C. A graphical method for exploring heterogeneity in meta-analyses: application to a meta-analysis of 65 trials. Statistics in Medicine. 2002;21(18):2641–52. doi: 10.1002/sim.1221. PubMed PMID: WOS:000178003300003.

38. DerSimonian R, Laird N. Meta-analysis in clinical trials. Control Clin Trials.1986;7(3):177–88. Epub 1986/09/01. doi: 10.1016/0197-2456(86)90046-2. PubMed PMID: 3802833.

39. Egger M, Davey Smith G, Schneider M, Minder C. Bias in meta-analysis detected by a simple, graphical test. BMJ. 1997;315(7109):629–34. Epub 1997/10/06. doi: 10.1136/bmj.315.7109.629. PubMed PMID: 9310563; PubMed Central PMCID: PMCPMC2127453.

40. Tobias A. Assessing the influence of a single study in the meta-analysis estimate. Stata Tech Bull. 1999;47:15–7.

41. Schwarzer G, Carpenter JR, Rücker G. Meta-analysis with R: Springer; 2015.

42. Harrer M, Cuijpers P, Furukawa T, Ebert D. dmetar: Companion R Package For The Guide “ Doing Meta-Analysis in R”. 2019.

43. Guan HY, Yu NN, Wang H, Boswell M, Shi YJ, Rozelle S, et al. Impact of various types of near work and time spent outdoors at different times of day on visual acuity and refractive error among Chinese school-going children. Plos One. 2019;14(4). doi: ARTN e0215827 10.1371/journal.pone.0215827. PubMed PMID: WOS:000465846200021.

44. Alharbi MT, Alem AA, Alrizgi HA, Abualnassr KA, Abualnassr FA, Alshencieti AM, et al. Impact of Smartphones on Eye Health among Health Sciences Students in Taibah University, Saudi Arabia. Indo Am J Pharm Sci. 2019;6(2):3523–30. doi: 10.5281/zenodo.2561230. PubMed PMID: WOS:000458607000103.

45. Küçer N. Some ocular symptoms experienced by users of mobile phones. Electromagn Biol Med. 2008;27(2):205–9. doi: 10.1080/15368370802072174. PubMed PMID: WOS:000256627200011.

46. Meo SA, Al-Drees AM. Mobile phone related-hazards and subjective hearing and vision symptoms in the Saudi population. Int J Occup Med Environ Health. 2005;18(1):53–7. Epub 2005/08/02. PubMed PMID: 16052891.

47. Merrie YA, Tegegne MM, Munaw MB, Alemu HW. Prevalence And Associated Factors Of Visual Impairment Among School-Age Children In Bahir Dar City, Northwest Ethiopia. Clin Optom. 2019;11:135–43. doi: 10.2147/Opto.S213047. PubMed PMID: WOS:000495435400001.

48. Lee D, Hong S, Jung S, Lee K, Lee G. The Effects of Viewing Smart Devices on Static Balance, Oculomotor Function, and Dizziness in Healthy Adults. Med Sci Monitor. 2019;25:8055–60. doi: 10.12659/Msm.915284. PubMed PMID: WOS:000492869400002.

49. Zylbermann R, Landau D, Berson D. The influence of study habits on myopia in Jewish teenagers. J Pediatr Ophthalmol Strabismus. 1993;30(5):319–22. Epub 1993/09/01. PubMed PMID: 8254449.

50. Lin LL, Shih YF, Hsiao CK, Chen CJ. Prevalence of myopia in Taiwanese schoolchildren: 1983 to 2000. Ann Acad Med Singapore. 2004;33(1):27–33. Epub 2004/03/11. PubMed PMID:15008558.

51. Morgan I, Rose K. How genetic is school myopia? Prog Retin Eye Res. 2005;24(1):1–381. Epub 2004/11/24. doi: 10.1016/j.preteyeres.2004.06.004. PubMed PMID: 15555525.

52. Morgan IG, French AN, Ashby RS, Guo X, Ding X, He M, et al. The epidemics of myopia: Aetiology and prevention. Prog Retin Eye Res. 2018;62:134–49. Epub 2017/09/28. doi: 10.1016/j.preteyeres.2017.09.004. PubMed PMID: 28951126.

53. 53. Sensaki S, Sabanayagam C, Verkicharla PK, Awodele A, Tan KH>, Chia A, et al. An Ecologic Study of Trends in the Prevalence of Myopia in Chinese Adults in Singapore Born from the 1920s to 1980s. Ann Acad Med Singapore. 2017;46(6):229–36. Epub 2017/07/25. PubMed PMID: 28733687.

54. Morgan IG, Rose KA. Myopia: is the nature-nurture debate finally over? Clin Exp Optom. 2019;102(1):3–17. Epub 2018/11/01. doi: 10.1111/cxo.12845. PubMed PMID: 30380590.

55. Hansen MH, Laigaard PP, Olsen EM, Skovgaard AM, Larsen M, Kessel L, et al. Low physical activity and higher use of screen devices are associated with myopia at the age of 16–17 years in the CCC2000 Eye Study. Acta Ophthalmol. 2020;98(3):315–21. Epub 2019/09/11. doi: 10.1111/aos.14242. PubMed PMID: 31502414.

56. Gopinath B, Hardy LL, Baur LA, Teber E, Mitchell P. Influence of parental history of hypertension on screen time and physical activity in young offspring. J Hypertens. 2012;30(2):336–41. Epub 2011/12/20. doi: 10.1097/HJH.0b013e32834ea4361. PubMed PMID: 22179085.

57. Harrington SC, Stack J, O’Dwyer V. Risk factors associated with myopia in schoolchildren in Ireland. Br J Ophthalmol. 2019;103(12):1803–9. Epub 2019/02/13. doi: 10.1136/bjophthalmol-2018-313325. PubMed PMID: 30745305.

58. Lanca C, Saw SM. The association between digital screen time and myopia: A systematic review. Ophthal Physl Opt. 2020;40(2):216–29. doi: 10.1111/opo.12657. PubMed PMID: WOS:000506849300001.

59. Yang GY, Huang LH, Schmid KL, Li CG, Chen JY, He GH, et al. Associations Between Screen Exposure in Early Life and Myopia amongst Chinese Preschoolers. Int J Env Res Pub He. 2020;17(3). doi: ARTN 1056 10.3390/ijerph17031056. PubMed PMID: WOS:000517783300381.

60. Toh SH, Coenen P, Howie EK, Smith AJ, Mukherjee S, Mackey DA, et al. A prospective longitudinal study of mobile touch screen device use and musculoskeletal symptoms and visual health in adolescents. Appl Ergon. 2020;85. doi: ARTN 103028 10.1016/j.apergo.2019.103028. PubMed PMID: WOS:000526996900002.

61. Enthoven CA, Tideman JWL, Polling JR, Yang-Huang J, Raat H, Klaver CCW. The impact of computer use on myopia development in childhood: The Generation R study. Prev Med.2020;132:105988. Epub 2020/01/19. doi: 10.1016/j.ypmed.2020.105988. PubMed PMID: 31954142.

62. Jones-Jordan LA, Mitchell GL, Cotter SA, Kleinstein RN, Manny RE, Mutti DO, et al. Visual activity before and after the onset of juvenile myopia. Invest Ophthalmol Vis Sci. 2011;52(3):1841–50. Epub 2010/10/12. doi: 10.1167/iovs.09-4997. PubMed PMID: 20926821; PubMed Central PMCID: PMCPMC3101696.

63. Wu PC, Tsai CL, Wu HL, Yang YH, Kuo HK. Outdoor activity during class recess reduces myopia onset and progression in school children. Ophthalmology. 2013;120(5):1080–5. Epub 2013/03/07. doi: 10.1016/j.ophtha.2012.11.009. PubMed PMID: 23462271.

64. Ramamurthy D, Lin Chua SY, Saw SM. A review of environmental risk factors for myopia during early life, childhood and adolescence. Clin Exp Optom. 2015;98(6):497–506. Epub 2015/10/27. doi: 10.1111/cxo.12346. PubMed PMID: 26497977.

65. Dirani M, Crowston JG, Wong TY. From reading books to increased smart device screen time. Br J Ophthalmol. 2019;103(1):1–2. Epub 2018/11/08. doi: 10.1136/bjophthalmol-2018-313295. PubMed PMID: 30401675.

66. McCloskey M, Johnson SL, Benz C, Thompson DA, Chamberlin B, Clark L, et al. Parent Perceptions of Mobile Device Use Among Preschool-Aged Children in Rural Head Start Centers. J Nutr Educ Behav. 2018;50(1):83–9 e1. Epub 2017/10/17. doi: 10.1016/j.jneb.2017.03.006. PubMed PMID: 29031581.

67. Wilkie HJ, Standage M, Gillison FB, Cumming SP, Katzmarzyk PT. The home electronic media environment and parental safety concerns: relationships with outdoor time after school and over the weekend among 9-11 year old children. BMC Public Health. 2018;18(1):456. Epub 2018/04/07. doi: 10.1186/s12889-018-5382-0. PubMed PMID: 29621981; PubMed Central PMCID: PMCPMC5887248.

68. Hardell L, Sage C. Biological effects from electromagnetic field exposure and public exposure standards. Biomed Pharmacother. 2008;62(2):104–9. Epub 2008/02/05. doi: 10.1016/j.biopha.2007.12.004. PubMed PMID: 18242044.

69. Kheifets L, Repacholi M, Saunders R, van Deventer E. The sensitivity of children to electromagnetic fields. Pediatrics. 2005;116(2):e303–13. Epub 2005/08/03. doi: 10.1542/peds.2004-2541. PubMed PMID: 16061584.

70. Balci M, Devrim E, Durak I. Effects of mobile phones on oxidant/antioxidant balance in cornea and lens of rats. Curr Eye Res. 2007;32(1):21–5. Epub 2007/03/17. doi: 10.1080/02713680601114948. PubMed PMID: 17364731.

71. Akar A, Karayigit MO, Bolat D, Gultiken ME, Yarim M, Castellani G. Effects of low level electromagnetic field exposure at 2.45 GHz on rat cornea. Int J Radiat Biol. 2013;89(4):243–9. Epub 2012/12/05. doi: 10.3109/09553002.2013.754557. PubMed PMID: 23206266.

72. Lixia S, Yao K, Kaijun W, Deqiang L, Huajun H, Xiangwei G, et al. Effects of 1.8 GHz radiofrequency field on DNA damage and expression of heat shock protein 70 in human lens epithelial cells. Mutat Res. 2006;602(1–2):135–42. Epub 2006/10/03. doi: 10.1016/j.mrfmmm.2006.08.010. PubMed PMID: 17011595.

73. Leitgeb N. Mobile phones: are children at higher risk? Wien Med Wochenschr. 2008;158(1–2):36–41. Epub 2008/02/21. doi:10.1007/s10354-007-0447-1. PubMed PMID: 18286248.

74. Khurana VG, Teo C, Kundi M, Hardell L, Carlberg M. Cell phones and brain tumors: a review including the long-term epidemiologic data. Surg Neurol. 2009;72(3):205–14; discussion 14–5. Epub 2009/03/31. doi: 10.1016/j.surneu.2009.01.019. PubMed PMID: 19328536.

75. Elder JA. Ocular effects of radiofrequency energy. Bioelectromagnetics. 2003;Suppl 6:S148–61. Epub 2003/11/25. doi: 10.1002/bem.10117. PubMed PMID: 14628311.

76. Yao K, Wang KJ, Sun ZH, Tan J, Xu W, Zhu LJ, et al. Low power microwave radiation inhibits the proliferation of rabbit lens epithelial cells by upregulating P27Kip1 expression. Mol Vis. 2004;10:138–43. Epub 2004/03/03. PubMed PMID: 14990889.

77. Challis LJ. Mechanisms for interaction between RF fields and biological tissue. Bioelectromagnetics. 2005;Suppl 7:S98–S106. Epub 2005/06/03. doi: 10.1002/bem.20119. PubMed PMID: 15931683.

78. Jones LA, Sinnott LT, Mutti DO, Mitchell GL, Moeschberger ML, Zadnik K. Parental history of myopia, sports and outdoor activities, and future myopia. Invest Ophthalmol Vis Sci. 2007;48(8):3524–32. Epub 2007/07/27. doi: 10.1167/iovs.06-1118. PubMed PMID: 17652719; PubMed Central PMCID: PMCPMC2871403.

79. Rose KA, Morgan IG, Ip J, Kifley A, Huynh S, Smith W, et al. Outdoor activity reduces the prevalence of myopia in children. Ophthalmology. 2008;115(8):1279–85. Epub 2008/02/26. doi: 10.1016/j.ophtha.2007.12.019. PubMed PMID: 18294691.

